# IS THE INVERSE ASSOCIATION BETWEEN PARKINSON’S DISEASE AND SMOKING DUE TO CONFOUNDING?

**DOI:** 10.1101/2020.09.20.20198390

**Authors:** Susan M. Davis

## Abstract

Parkinson’s disease (PD) is inversely associated with smoking. Whether this association is due to a causal relationship or to confounding by a covariate of smoking is still debated.

This study included populations of the United States and ten U.S. states from 2004 to 2016. The ten U.S. states included the five states with the highest PD incidence rates in 2017 (Vermont, Maine, Kansas, Missouri, Colorado) and the five states with the lowest PD incidence rates in 2017 (Arkansas, Mississippi, South Dakota, Nebraska, Alabama). The study used scatter plots to explore the association between PD incidence and smoking and the association between PD incidence and a covariate of smoking, lower endoscopy utilization.

The results indicate that there is a weaker association between PD and smoking when compared to the association between PD and lower endoscopy utilization. For PD incidence verses current smoking, the coefficient of determination (R^2^) for the United States was 0.813 and ranged from a low of 0.000 for Arkansas to 0.798 for Colorado (average R^2^ for the ten states was 0.391). For PD incidence verses lower endoscopy utilization, the R^2^ for the United States was 0.995 and ranged from a low of 0.871 for South Dakota to 0.997 for Colorado (average R^2^ for the ten states was 0.959).

The results suggest that the inverse association between PD incidence and smoking is confounded by the direct association between PD and lower endoscopy utilization. Further investigation of a possible relationship between PD incidence and lower endoscopy utilization is warranted.

## INTRODUCTION

Parkinson’s disease (PD) incidence rates have been increasing [1]. There is no known cause for most cases of PD, and they are thought to result from a complex combination of many factors, including industrialization, environmental toxins, lifestyle, genetics, and age [2-3].

PD is related to smoking, with current smokers having a lower risk for PD when compared to never smokers [2-4]. Of eighty-four risk factors explored by the Global Burden of Diseases, Injuries, and Risk Factors (GBD), smoking was the only risk factor deemed to have a relationship with PD [2]. The inverse relationship between PD and smoking may be casual or it may be due to confounding by a covariate of smoking.

One covariate of smoking is colorectal cancer (CRC) screening. Several studies have found that current smokers are less likely to participate in CRC screening compared to never smokers [5-9]. In the United States, the most commonly used CRC screening test is colonoscopy [10]. Compared with never smokers, current smokers are less likely to ever have received a colonoscopy [5].

Colonoscopy is a lower endoscopic procedure that examines the entire colon, and precancerous polyps can be removed during the procedure, thereby reducing CRC risk [11-12]. Another lower endoscopic procedure used for CRC screening is sigmoidoscopy, which is similar to colonoscopy, but examines only a portion of the colon [12-13].

## METHODS

The present study comprises scatter plots of PD incidence versus current smoking and PD incidence versus lower endoscopy utilization for different populations.

This study included populations of the United States (all 50 states and the District of Columbia) and ten U.S. states from 2004 to 2016. The ten U.S. states included the five states with the highest PD incidence rates in 2017 (Vermont, Maine, Kansas, Missouri, Colorado) and the five states with the lowest PD incidence rates in 2017 (Arkansas, Mississippi, South Dakota, Nebraska, Alabama).

PD incidence rates (age standardized, new cases per 100,000) were obtained from the Institute for Health Metrics and Evaluation (IHME)’s Global Burden of Diseases, Injuries, and Risk Factors (GBD) Compare visualization system (available at https://vizhub.healthdata.org/gbd-compare) [14]. Methodology, sample size, and 95% confidence intervals may be obtained from the IHME.

Rates of current smoking (% aged ≥18 who currently smoke cigarettes) and rates of lower endoscopy utilization (% *a*ged ≥50 who have had a sigmoidoscopy or colonoscopy) were obtained from the Centers for Disease Control and Prevention (CDC)’s Behavioral Risk Factor Surveillance System (BRFSS) Web Enabled Analysis Tool (WEAT) (available at https://www.cdc.gov/brfss/data_tools.htm) [15]. Methodology, sample size, and 95% confidence intervals may be obtained from the CDC.

The scatter plots, trendline equations and coefficients of determination (R^2^) were generated using Microsoft Excel.

## RESULTS

Scatter plots of PD verses smoking and PD verses lower endoscopy are shown in Figures 1 and 2, respectively. The data plotted in Figures 1 and 2, the trendline equations, and the R^2^ values are shown in Table 1.

**Table 1:**
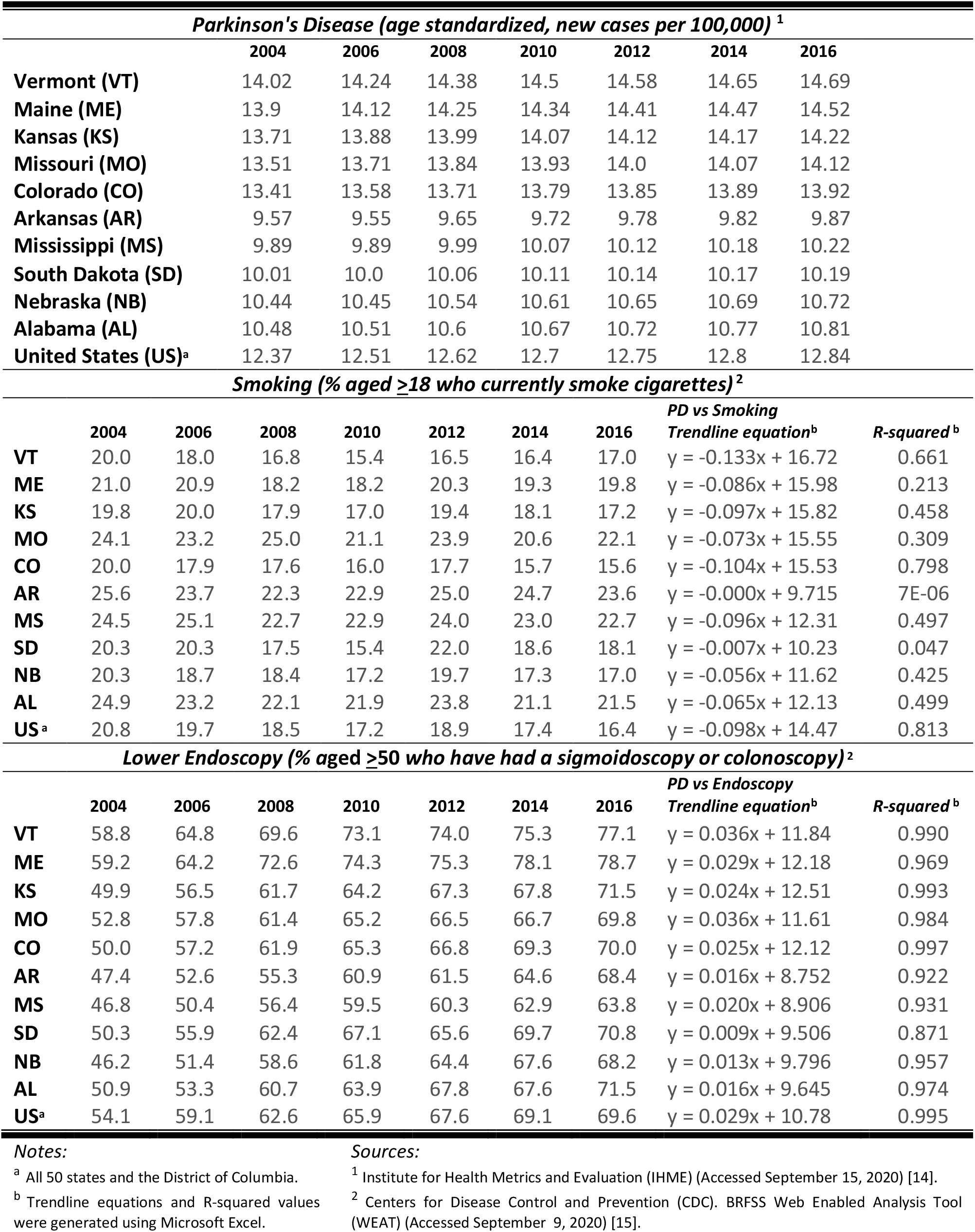
Data plotted in Figures 1 and 2.

| Parkinson's Disease (age standardized, new cases per 100,000) <sup>1</sup> |  |  |  |  |  |  |  |  |  |
| --- | --- | --- | --- | --- | --- | --- | --- | --- | --- |
|  | 2004 | 2006 | 2008 | 2010 | 2012 | 2014 | 2016 |  |  |
| Vermont (VT) | 14.02 | 14.24 | 14.38 | 14.5 | 14.58 | 14.65 | 14.69 |  |  |
| Maine (ME) | 13.9 | 14.12 | 14.25 | 14.34 | 14.41 | 14.47 | 14.52 |  |  |
| Kansas (KS) | 13.71 | 13.88 | 13.99 | 14.07 | 14.12 | 14.17 | 14.22 |  |  |
| Missouri (MO) | 13.51 | 13.71 | 13.84 | 13.93 | 14.0 | 14.07 | 14.12 |  |  |
| Colorado (CO) | 13.41 | 13.58 | 13.71 | 13.79 | 13.85 | 13.89 | 13.92 |  |  |
| Arkansas (AR) | 9.57 | 9.55 | 9.65 | 9.72 | 9.78 | 9.82 | 9.87 |  |  |
| Mississippi (MS) | 9.89 | 9.89 | 9.99 | 10.07 | 10.12 | 10.18 | 10.22 |  |  |
| South Dakota (SD) | 10.01 | 10.0 | 10.06 | 10.11 | 10.14 | 10.17 | 10.19 |  |  |
| Nebraska (NB) | 10.44 | 10.45 | 10.54 | 10.61 | 10.65 | 10.69 | 10.72 |  |  |
| Alabama (AL) | 10.48 | 10.51 | 10.6 | 10.67 | 10.72 | 10.77 | 10.81 |  |  |
| United States (US)<sup>a</sup> | 12.37 | 12.51 | 12.62 | 12.7 | 12.75 | 12.8 | 12.84 |  |  |
| Smoking (% aged ≥18 who currently smoke cigarettes)<sup>2</sup> |  |  |  |  |  |  |  |  |  |
|  | 2004 | 2006 | 2008 | 2010 | 2012 | 2014 | 2016 | PD vs Smoking<br>Trendline equation<sup>b</sup> | R-squared<sup>b</sup> |
| VT | 20.0 | 18.0 | 16.8 | 15.4 | 16.5 | 16.4 | 17.0 | y = -0.133x + 16.72 | 0.661 |
| ME | 21.0 | 20.9 | 18.2 | 18.2 | 20.3 | 19.3 | 19.8 | y = -0.086x + 15.98 | 0.213 |
| KS | 19.8 | 20.0 | 17.9 | 17.0 | 19.4 | 18.1 | 17.2 | y = -0.097x + 15.82 | 0.458 |
| MO | 24.1 | 23.2 | 25.0 | 21.1 | 23.9 | 20.6 | 22.1 | y = -0.073x + 15.55 | 0.309 |
| CO | 20.0 | 17.9 | 17.6 | 16.0 | 17.7 | 15.7 | 15.6 | y = -0.104x + 15.53 | 0.798 |
| AR | 25.6 | 23.7 | 22.3 | 22.9 | 25.0 | 24.7 | 23.6 | y = -0.000x + 9.715 | 7E-06 |
| MS | 24.5 | 25.1 | 22.7 | 22.9 | 24.0 | 23.0 | 22.7 | y = -0.096x + 12.31 | 0.497 |
| SD | 20.3 | 20.3 | 17.5 | 15.4 | 22.0 | 18.6 | 18.1 | y = -0.007x + 10.23 | 0.047 |
| NB | 20.3 | 18.7 | 18.4 | 17.2 | 19.7 | 17.3 | 17.0 | y = -0.056x + 11.62 | 0.425 |
| AL | 24.9 | 23.2 | 22.1 | 21.9 | 23.8 | 21.1 | 21.5 | y = -0.065x + 12.13 | 0.499 |
| US<sup>a</sup> | 20.8 | 19.7 | 18.5 | 17.2 | 18.9 | 17.4 | 16.4 | y = -0.098x + 14.47 | 0.813 |
| Lower Endoscopy (% aged ≥50 who have had a sigmoidoscopy or colonoscopy)<sup>2</sup> |  |  |  |  |  |  |  |  |  |
|  | 2004 | 2006 | 2008 | 2010 | 2012 | 2014 | 2016 | PD vs Endoscopy<br>Trendline equation<sup>b</sup> | R-squared<sup>b</sup> |
| VT | 58.8 | 64.8 | 69.6 | 73.1 | 74.0 | 75.3 | 77.1 | y = 0.036x + 11.84 | 0.990 |
| ME | 59.2 | 64.2 | 72.6 | 74.3 | 75.3 | 78.1 | 78.7 | y = 0.029x + 12.18 | 0.969 |
| KS | 49.9 | 56.5 | 61.7 | 64.2 | 67.3 | 67.8 | 71.5 | y = 0.024x + 12.51 | 0.993 |
| MO | 52.8 | 57.8 | 61.4 | 65.2 | 66.5 | 66.7 | 69.8 | y = 0.036x + 11.61 | 0.984 |
| CO | 50.0 | 57.2 | 61.9 | 65.3 | 66.8 | 69.3 | 70.0 | y = 0.025x + 12.12 | 0.997 |
| AR | 47.4 | 52.6 | 55.3 | 60.9 | 61.5 | 64.6 | 68.4 | y = 0.016x + 8.752 | 0.922 |
| MS | 46.8 | 50.4 | 56.4 | 59.5 | 60.3 | 62.9 | 63.8 | y = 0.020x + 8.906 | 0.931 |
| SD | 50.3 | 55.9 | 62.4 | 67.1 | 65.6 | 69.7 | 70.8 | y = 0.009x + 9.506 | 0.871 |
| NB | 46.2 | 51.4 | 58.6 | 61.8 | 64.4 | 67.6 | 68.2 | y = 0.013x + 9.796 | 0.957 |
| AL | 50.9 | 53.3 | 60.7 | 63.9 | 67.8 | 67.6 | 71.5 | y = 0.016x + 9.645 | 0.974 |
| US<sup>a</sup> | 54.1 | 59.1 | 62.6 | 65.9 | 67.6 | 69.1 | 69.6 | y = 0.029x + 10.78 | 0.995 |

**Notes:**<sup>a</sup> All 50 states and the District of Columbia.<sup>b</sup> Trendline equations and R-squared values were generated using Microsoft Excel.**Sources:**<sup>1</sup> Institute for Health Metrics and Evaluation (IHME) (Accessed September 15, 2020) [14].<sup>2</sup> Centers for Disease Control and Prevention (CDC). BRFSS Web Enabled Analysis Tool (WEAT) (Accessed September 9, 2020) [15].

**Figure 1:**
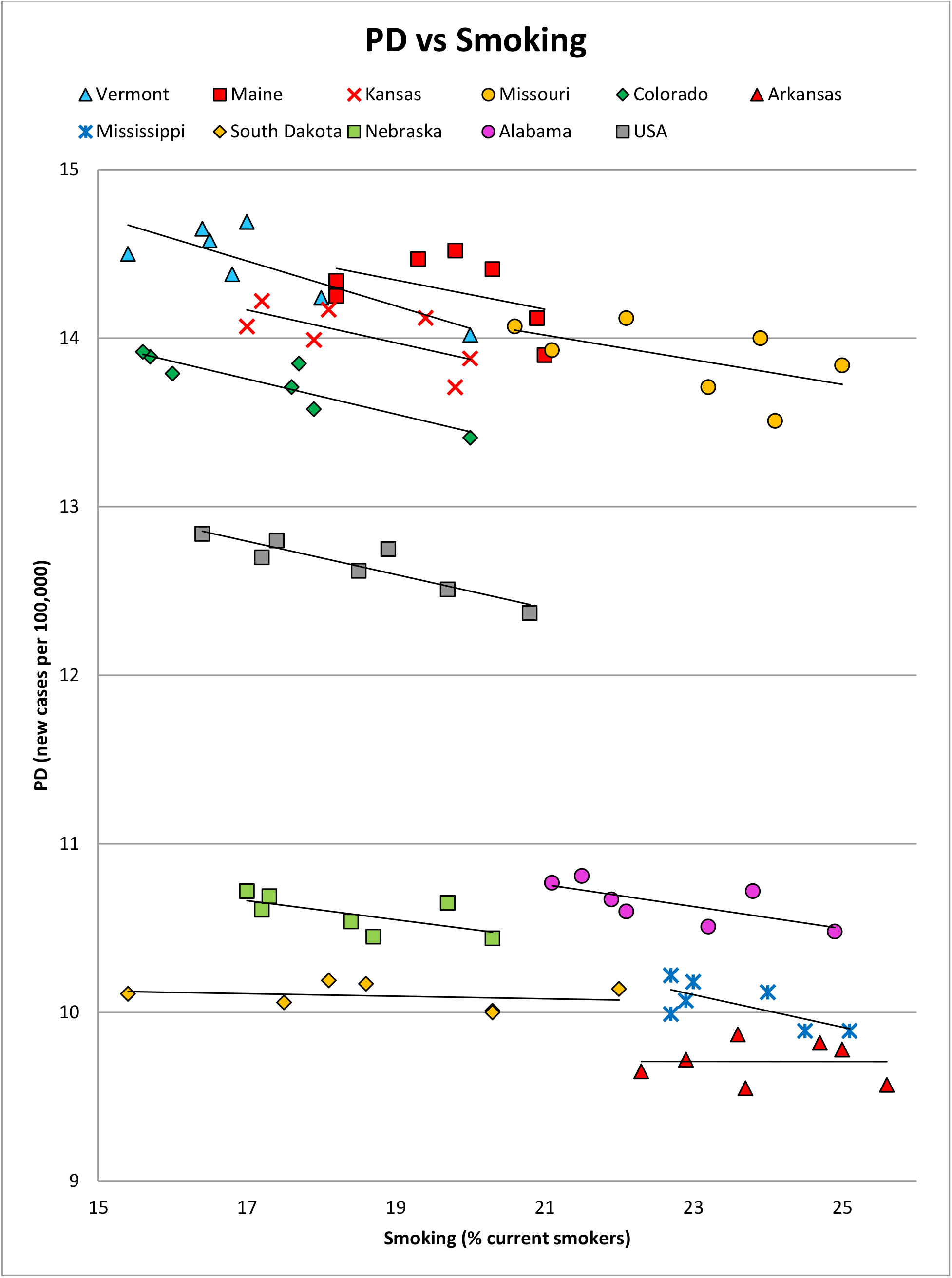
Parkinson’s disease incidence verses current smoking.

**Figure 2:**
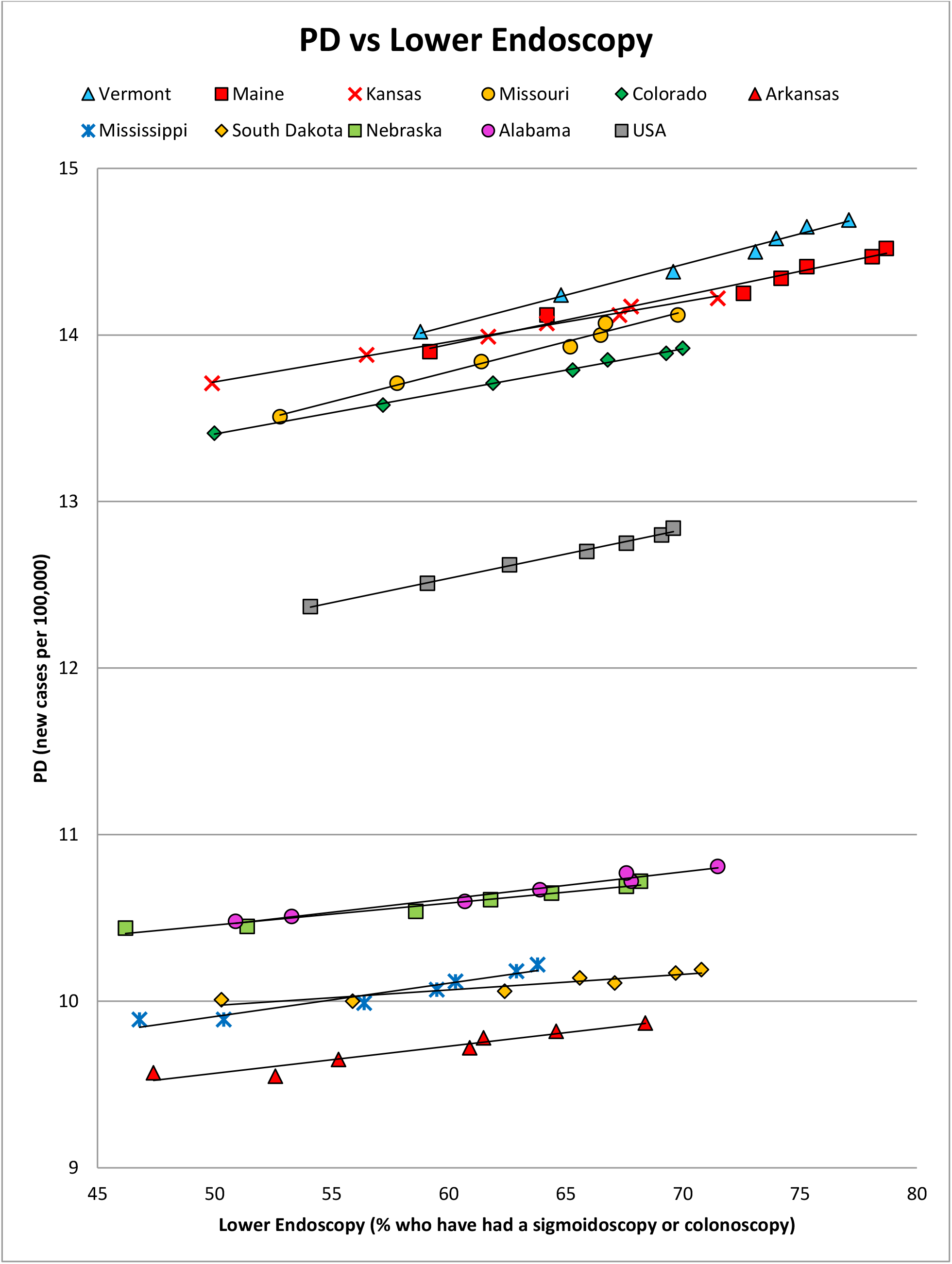
Parkinson’s disease incidence verses lower endoscopy usage.

As can be seen from the above scatter plots and table, there is a negative linear association between PD and smoking and a positive linear association between PD and lower endoscopy utilization. The correlation between PD and smoking is weaker than the correlation between PD and lower endscopy utilization for all 10 states and the U.S., as indicated by the R^2^ values.

## DISCUSSION

The results presented here indicate that there is a strong direct correlation between PD incidence and lower endoscopy utilization. It seems likely that the inverse association between PD and smoking is confounded by lower endoscopy utilization.

The inverse association between PD and smoking is not seen in ages ≥75 years [16]; and the U.S. Preventive Services Task Force (USPSTF) recommends CRC screening for ages 50-75 years [17], further suggesting confounding by lower endoscopy utilization.

Lower endoscopy utilization reduces CRC risk [11-13]. Persons with PD have a decreased risk of CRC when compared to persons without PD [18-20], although this decreased risk is not seen in persons diagnosed with PD at an age of ≥75 years [18]. Since the USPSTF recommends CRC screening for ages 50-75 years [17], it seems likely that the inverse association between PD and CRC risk is also confounded by lower endoscopy utilization.

I theorize that both the inverse association between PD and smoking and the inverse association between PD and CRC are confounded by the direct correlation between PD and lower endoscopy utilization. A biological explanation for a direct association between PD and lower endoscopy utilization is provided by the etiology of PD.

The present study can only conclude that there is a correlation between PD and lower endoscopy utilization at a population level; however, another study found that patients undergoing diagnostic colonoscopies may have an increased prevalence of clinically asymptomatic PD (prodromal PD) compared to the general elderly population [21], which suggests that patients undergoing colonoscopy may have an increased PD risk.

A limitation of the present study is that the characteristics of the population base may not have remained unchanged throughout the study period. Other limitations include the smoking and lower endoscopy percentages provided by the CDC, which are estimates based on the Behavioral Risk Factor Surveillance System (BRFSS), and the PD incidence rates provided by the IHME, which are also estimates. Another limitation is that covariates of lower endoscopy utilization cannot be excluded.

## CONCLUSIONS

The results indicate that the inverse association between PD and smoking is confounded by a direct association between PD and lower endoscopy utilization. Further investigation into the relationship between PD and lower endoscopy utilization is warranted and may provide a means for reducing PD incidence.

## Data Availability

Parkinson's disease incidence data is available from the Institute for Health Metrics and Evaluation (IHME)'s Global Burden of Diseases, Injuries, and Risk Factors (GBD) Compare visualization system (available at https://vizhub.healthdata.org/gbd-compare).
Current smoking and endoscopy usage data are available from the Centers for Disease Control and Prevention (CDC)'s Behavioral Risk Factor Surveillance System (BRFSS) Web Enabled Analysis Tool (WEAT) (available at https://www.cdc.gov/brfss/data_tools.htm). 

https://www.cdc.gov/brfss/data_tools.htm

## ACKNOWLEDGMENTS

The insight of Emily E. Davis was instrumental in the conception of this study.

